# Diagnostic accuracy of five mpox lateral flow assays for antigen detection, the Democratic Republic of the Congo and Switzerland

**DOI:** 10.64898/2026.01.28.26345023

**Authors:** Hugo Kavunga-Membo, Hanesh Fru Chi, Devy M. Emperador, Hugues Mirimo, Elie Ishara-Nshombo, Marithe Mukoka, Raphael Lumembe, Michael Otieno, Mireille Muloki, Jean-Claude Makangara-Cigolo, Eddy Kinganda-Lusamaki, Tony Wawina-Bokalanga, Camille Escafadal, Kenneth Adea, Ana Hoxha, Rosamund F. Lewis, Nicksy Gumede, Olga Ntumba-Tshitenge, Richard Fotsing, Isabella Eckerle, Daniel Mukadi-Bamuleka, Placide Mbala-Kigenbeni, Emmanuel Agogo, Lorenzo Subissi

## Abstract

**Background:** Mpox has spread to 35 countries in Africa, yet many face challenges achieving nationwide PCR-based testing due to cost and limited access in remote and rural areas. Point-of-care antigen-based rapid diagnostic tests (AgRDTs) may help improve diagnostic access.

**Objectives:** To evaluate the diagnostic accuracy of five mpox AgRDTs manufactured by Beijing Hotgen Biotech (China), Contipharma (Belgium), Hangzhou Testsea Biotechnology (China), Guangdong Wesail Biotech (China), and NG Biotech (France).

**Methods:** Diagnostic accuracy was assessed using 190 lesion swabs from suspected mpox cases in the Democratic Republic of the Congo, with results compared against the RADI FAST Mpox PCR assay (KH Medical, South Korea). Analytical sensitivity was evaluated using a clade Ib MPXV isolate from the WHO Biohub held by the Geneva University Hospitals, Switzerland.

**Results:** The best-performing assay, the Monkeypox Virus Ag Test Kit (Guangdong Wesail Biotech), demonstrated a sensitivity of 77.3% (95% CI: 68.0–84.5) and specificity of 93.5% (95% CI: 86.6–97.0). The Hangzhou Testsea assay showed comparable performance (sensitivity 72.2%, specificity 93.5%). Beijing Hotgen and Contipharma assays exhibited moderate sensitivity (59.8% and 50.5%, respectively) with high specificity (96.8% and 95.7% respectively), while the NG Biotech assay showed the lowest sensitivity (39.2%) but similarly high specificity (96.8%). Analytical testing revealed no major differences across assays, though Guangdong Wesail demonstrated the highest analytical sensitivity, detecting clade Ib virus at a Ct of 28.3.

**Conclusion:** Several AgRDTs show high positive predictive values for mpox screening in high-prevalence settings, where positive test results may support confirmation but negative results cannot rule out infection. Further multicentre prospective studies are needed to define appropriate use cases as countries transition to sustainable mpox control.

**Research in context:** *Evidence before this study:* Access to decentralized mpox diagnostics remains limited in many African countries due to the cost, infrastructure, and turnaround time associated with PCR, particularly in remote settings. Antigen-based rapid diagnostic tests (AgRDTs) could expand access to point-of-care testing, but data supporting their clinical performance has been limited. We searched PubMed and medRxiv on Dec 31, 2025, using the following search string: (mpox OR monkeypox) AND (diagnostic* OR point of care OR lateral flow assay OR rapid antigen test OR AgRDT). Early evaluations of mpox AgRDTs reported very low sensitivity. A large prospective, multicentre field study in Uganda and the Democratic Republic of the Congo (DRC) evaluating a single mpox AgRDT for research use only using lesion swabs reported an overall sensitivity of 70.4%, with substantial variation by country (81.9% in Uganda vs 55.1% in DR Congo), age, and viral load, and specificity of 89.3%. Independent comparative evaluations of multiple commercially available AgRDTs with regulatory approval—particularly in clade I–endemic African settings—have been lacking.

*Added value of this study:* This study provides the first head-to-head clinical and analytical evaluation of five commercially available mpox AgRDTs using skin or mucosal lesion swab specimens from suspected cases in DRC. By assessing all assays against a molecular reference test under identical conditions and incorporating analytical testing with a clade Ib reference virus isolate, we demonstrate substantial variability in performance between manufacturers tests kits. Two assays achieved sensitivity against PCR of above 70% with moderate specificity (93.5%), comparable to or exceeding results reported in prior field studies, while others showed moderate to poor sensitivity. Analytical testing identified meaningful differences in detection limits, with the best-performing assay detecting clade Ib virus at viral loads corresponding to a Ct value of 28.3. These findings provide critical data to inform further field-based validation projects, assay selection and procurement decisions for use of such AgRDTs in endemic settings.

*Implications of all the available evidence:* Across studies, mpox AgRDTs consistently demonstrate high specificity but variable and often insufficient sensitivity to function as standalone diagnostic tools. Prior studies have provided evidence that sensitivity is highest for samples with Ct values 25 or lower, and the present comparative evaluation show that positive AgRDT results may be able to reliably confirm mpox infection in high-prevalence settings, whereas negative results cannot safely exclude disease. The heterogeneity in performance between assays underscores the need for field-based independent evaluations of most promising tests before their large-scale deployment, and cautions against treating AgRDTs as a uniform diagnostic test category. As countries transition from emergency response to sustainable mpox control, AgRDTs may have a complementary role in decentralized screening, outbreak confirmation, and triage, provided confirmatory PCR remains available. Further multi-centre prospective studies and continued assay optimization are essential to ensure antigen-based diagnostics can be used to meaningfully strengthen mpox surveillance, preparedness and outbreak response.

## Introduction

The monkeypox virus (MPXV) is a double-stranded DNA virus belonging to the *Poxviridae* family, *Orthopoxvirus* genus—a group that also includes vaccinia, cowpox, and variola viruses.^1^ Orthopoxviruses are large oval-shaped viruses with almost 200 kb genomes coding for over 200 genes.^2^ MPXV was first identified in 1958 during an outbreak among captive cynomolgus macaques (*Macaca fascicularis*) transported from Singapore to Denmark.^3^ More than a decade later, in 1970, the first human case of mpox, the disease caused by MPXV, was documented in the Democratic Republic of the Congo (DRC).^4^

In most circumstances, such as new outbreaks, ongoing surveillance and in consideration of differential diagnoses, suspected mpox should be laboratory-confirmed, with nucleic acid amplification tests (NAAT) currently serving as the gold standard assay for mpox.^5^ However, PCR-based assays require specialized instruments, DNA extraction, and skilled laboratory personnel, factors that hinder accessibility in low- and middle-income countries (LMICs). Consequently, numerous suspected cases remain unconfirmed due to limited diagnostic infrastructure, fragile healthcare systems, and sociopolitical instability, delaying outbreak confirmation and response.

Since 2022, when the first Public Health Emergency of International Concern (PHEIC) was declared by WHO, multiple virus strains have emerged, causing substantial outbreaks with major public health impact across the world. MPXV clade IIb/sh2017, after its initial detection in 2017 in Nigeria, caused mpox isolated cases or larger outbreaks in over 110 countries or territories from May 2022 onwards, with most cases among men who have sex with men (MSM).^6^ In 2023 and 2024, clade Ib/sh2023 and clade Ia/sh2024 emerged in South Kivu and Kinshasa provinces (DRC) respectively, and both affected sex workers, their clients as well as resulting in secondary cases in households, including among children.^7,8^ While Clade Ib/sh2023 spread to 35 African countries and still causes community transmission in many of them, clade Ia/sh2024 was found in the DRC, and subsequently exported to Ireland, Turkiye and China.^9^

Between January 2024 and May 2025, the epidemiology of mpox in the Democratic Republic of the Congo was marked by the emergence and geographic expansion of clade Ib/sh2023, which originated in South Kivu Province and subsequently spread to most of the 26 provinces in the country. Concurrently, clade Ia transmission persisted in historically endemic areas, with both clades co-circulating in Kinshasa.^8^ On 14 August 2024, the WHO declared a second PHEIC following a renewed rise in mpox clade I cases in Africa, with potential for international spread. This triggered opening of the WHO Emergency use listing (EUL) for mpox in vitro diagnostics (IVD),^10^ and renewed urgency for accelerated innovation in rapid diagnostic technologies, including in NAAT platforms designed for near-patient testing as well as antigen-based lateral flow assays (AgRDTs).

Near-patient POC NAAT systems, which were listed in the WHO EUL for mpox IVD shortly after its opening and used since the second PHEIC in a number of African countries, have been critical to decentralized testing strategies.^5,11,12^ For example, in the DRC, at the end of 2023, only two laboratories were testing for mpox (INRB Goma and INRB Kinshasa) and the proportion of suspected cases tested was as low as 9%.^13^ By August 2025, thanks to the roll-out of near-patient NAAT systems which had benefitted from WHO EUL, the number of laboratories able to test for mpox increased to 29 and testing of suspected mpox cases reached 60%. However, at least 40% of suspected mpox cases remained without access to testing. In addition, testing with near-patient NAAT systems remains costly at approximately USD 20.00 per test, raising valid questions about the sustainability of their use outside emergency settings.

Accurate and rapid laboratory confirmation of infectious diseases remain central to rapid outbreak response and to effective infection control, enabling prompt isolation and optimal clinical management. In July 2023, WHO published target product profiles (TPPs) for tests to diagnose mpox, which included tests used as an aid to diagnosis by detecting orthopoxvirus (OPXV) antigens, which are amenable to decentralized use, including in the community. This TPP set minimal criteria for clinical sensitivity and specificity to ≥80% and ≥97%, respectively. ^14^ Antigen rapid diagnostic tests have enormous potential in supporting rapid, simple and affordable confirmation and response to MPXV outbreaks especially in decentralized settings. To date, few studies reported diagnostic accuracy of AgRDTs: one commercial lateral flow device (LFD, Tetracore Orthopox Biothreat Alert) reproducibly detected laboratory grown vaccinia and mpox viruses containing 10^7^ pfu/ml, and identified samples containing 10^6^ pfu/ml in 4 of 7 independent experiments; it had promising sensitivity of 80% (9/11) and specificity of 90% (10/11) in a clinical evaluation, but those samples had low cycle threshold (Ct) values between 15-22.^15^ Another study assessed an in house LFD, with a sensitivity 3 × 10^5^ pfu/mL, which was deemed sufficient for detection of virus in lesion swabs.^16^ Finally, another study observed very poor sensitivity despite high specificity for 3 commercially available AgRDTs using stored lesion samples in liquid buffer.^17^ Finally, a field-based prospective multicentre diagnostic accuracy study evaluated the NG-Test Monkeypox antigen RDT (NG Biotech, Guipry-Messac, France) and found a sensitivity of 70·4% (293 of 416 [95% CI 65·9-74·6]) and specificity of 89·3% (201 of 225 [95%CI 84·6-92·7]), showing the feasibility of field use of such tools, including in conflict zones. ^18^ In another study, clinical samples collected from MPXV patients showed a specificity of 100 % and overall sensitivity of 40%, with a sensitivity a high as 87 % at a qPCR cycle threshold of 25 or lower. ^19^ However, none of these studies met the accuracy benchmarks outlined in the WHO target product profiles, and the suitability of AgRDTs for use in clinical settings and as an adjunct to surveillance for outbreak confirmation and monitoring therefore remains uncertain. Therefore, independent performance data on emerging AgRDTs are needed to inform their use. In this study, we evaluated the diagnostic performance of five AgRDTs against a reference test for the detection of MPXV in lesion material.

## Methods

### Test selection

Tests were selected based on previous analytical data (and clinical data, if available) from AgRDT manufacturers through a request for proposal from FIND,^20^ complemented by advice from the WHO Collaborating Centre for Smallpox and other poxvirus infections at the US Centers for Disease Control and Prevention. Selection criteria were: geographical reach of the AgRDT manufacturer; technology Readiness Level 9; CE marked/commercially available; clinical sensitivity at least 80% (lesion swab); clinical specificity at least 97% (lesion swab); no cross-reactivity with endogenous substances and other human non-orthopoxviruses; time to result less or equal to 30 minutes; minimal pre-test processing required; no additional laboratory equipment required; all necessary materials and reagents included in the kit; maximum price per test of 5 USD; available risk management report. Based on the above criteria, five AgRDT were shortlisted (Supplementary Table 1). For the tests for which information is available, the antigen used in the test is from the A29 protein.

### Clinical evaluation

#### Study design and participants

We conducted a retrospective diagnostic accuracy study using archived skin lesion specimens collected during a WHO-led prospective mpox transmission study in the Democratic Republic of Congo (DRC). The parent study aimed to characterize mpox transmission dynamics and is being conducted at multiple sites in the DRC. Participants provided informed consent for future use of biological samples.

The present study represents a secondary analysis designed to evaluate the diagnostic performance of multiple antigen-based rapid diagnostic tests (AgRDTs), with PCR as the reference standard.

#### Specimen collection, testing and bias mitigation

In the parent study, lesion swabs were prospectively collected in 11 health zones of Kinshasa province (i.e. Bumbu, Kalamu II, Kingabwa, Kinshasa, Kisenso, Kokolo, Limete, Lingwala, Masina II) from patients with suspected mpox. For each participant, six dry lesion swabs were obtained from three lesions (two swabs per lesion), preferentially sampling lesions with similar morphology from the same body region. Dry swabs were stored without viral transport medium (VTM); VTM containing guanidinium thiocyanate was avoided due to its reported interference with AgRDT sensitivity.^16^

Specimens were included if the following conditions were met: informed consent for future use was available, swabs were stored without viral transport medium (i.e., dry swabs), and reference PCR results (using RADI FAST Mpox Detection Kit) confirming mpox positivity or negativity were available. Specimens were excluded if the collection date was unknown or prior to 2020, or if they had undergone more than three freeze–thaw cycles.

Because the parent study yielded a high mpox positivity among patients tested (76%), we supplemented the dataset with 37 additional PCR-negative wet-lesion swabs to improve the precision of specificity estimates. These samples had previously tested negative using the RADI FAST Mpox Detection Kit and were stored in phosphate-buffered saline at −80 °C until analysis

For each participant, one of the six collected dry swabs was rehydrated in phosphate-buffered saline (PBS) and used for reference testing with the RADI FAST Mpox Detection Kit (KH Medical, Pyeongtaek, South Korea), a WHO Emergency Use Listing–approved assay that can distinguish clade Ia and Ib.^10^ DNA was extracted from 200 µL of PBS using the RADI 32/48 Universal DNA/RNA Extraction Kit (KH Medical, Pyeongtaek, South Korea).

The remaining five dry swabs, stored at −80 °C, were thawed on the day of evaluation and rehydrated using the proprietary buffer supplied with each AgRDT, in accordance with the manufacturers’ instructions for use. Each lesion was used to evaluate one AgRDT, resulting in a total of five index tests per participant.

All the testing was done at INRB, Kinshasa, DRC. For each specimen, multiple index tests were performed. Each test was conducted and interpreted by an operator blinded to the results of all other tests. Testing was carried out in dedicated, physically separate areas to minimize interpretation bias. Reference PCR testing and index testing were performed and read by two different operators, all of whom were blinded to the results of other assays. A third operator was requested to interpret the result in case of discordant interpretation. Additional measures to minimize bias included blinding operators to the initial reference test result and ensuring that the same three laboratory personnel conducted testing throughout the study to reduce operator variability. Specimens were anonymized, and no participant-identifiable information was available to laboratory staff beyond PCR positivity status required for sample selection.

#### Ethics

Ethics approval was obtained from the National Ethics Committee of the DRC (ESP/CE/236/2024 and ESP/CE/11B/2025) and the WHO Regional Office for Africa Ethics Review Board (AFR/ERC/2024/9.6). The study was conducted in accordance with the Declaration of Helsinki, CIOMS guidelines, ISO 20916, ICH-GCP where applicable, and national regulatory requirements.

### Analytical evaluation

MPXV clade Ib was obtained from INRB, Kinshasa, DRC via the WHO BioHub System (MPXV, strain hMpxV/DRC-INRB/24MPX0203V/2024, clade Ib; ref 2024-WHO-LS-003). MPXV Clade Ib was propagated in Vero-E6 cells (ATCC CRL-1586) that were maintained in DMEM GlutaMAX I medium (Gibco, USA) supplemented with 10% FBS, 1× non-essential amino acids, and 1% penicillin-streptomycin. The supernatant was harvested to constitute a virus stock, and the cultured virus was fully sequenced (GenBank accession: PX049155.1). The viral stock was serially diluted in PBS with a dilution factor of 1:3, beginning at a concentration of 1.21×10^7^ DNA copies/mL. Multiple aliquots of each dilution were prepared and the viral load of each dilution was quantified by the PCR described by Scaramozzino *et al*. ^21^ targeting the open reading frame (ORF) A30L of orthopoxviruses. Viral DNA was extracted using the NucliSens easyMAG kit (BioMerieux), real-time PCR was performed using the *Taq* DNA Polymerase kit (Invitrogen) in a CFX96 Thermal Cycler (BIORAD). Quantification of DNA copies was done with in-house standards.

AgRDTs were performed according to the manufacturer’s instructions with the only difference that 200⍰μL of each virus dilution was added directly to the proprietary buffer tube instead of a swab specimen. PBS without virus was used as the negative control. Subsequently, the dilution/buffer mixture was applied to the sample pad of the Ag-RDT using the materials provided in the kit as recommended by manufacturer and following the instructions for use. The results were independently read by two individuals and interpreted according to the manufacturer’s instructions.

All work with infectious MPXV was conducted under BSL-3 conditions at the Geneva Centre for Emerging Viral Diseases, Geneva University Hospitals, Geneva, Switzerland.

### Statistical analysis

Diagnostic accuracy metrics were estimated by comparing the index test results against the RADI FAST Mpox detection kit results from lesion swabs from suspected cases of mpox. Point estimates of sensitivity and specificity were calculated as TP/(TP+FN) and TN/(TN+FP), respectively. Estimates were reported with 95% confidence intervals calculated using Wilson score method. Sample size calculations to estimate the sensitivity and specificity of each RDT were based on an expected sensitivity of 70% and specificity of 95% of the index test compared to the reference test. The study was powered to obtain a pooled sensitivity estimate within a 95% confidence interval with a probability (equivalent to power) of 80% to obtain a confidence interval of such width or less. The sample size (190 [97 MPXV positive and 93 MPXV negative] samples) was determined using a previously published formula [Equations 6.1 and 6.3].^22^

### Role of funding source

The funders of the study had no role in study design, data collection, data analysis, data interpretation, or writing of the report. The corresponding authors had full access to all the data in the study and had final responsibility for the decision to submit for publication.

## Results

### Clinical evaluation

A total of 918 lesion swabs from 153 patients were collected during a WHO field study of mpox transmission in the DRC, and a total of 802 lesions from 190 patients were tested using five AgRDTs (Figure 1).

**Figure 1:**
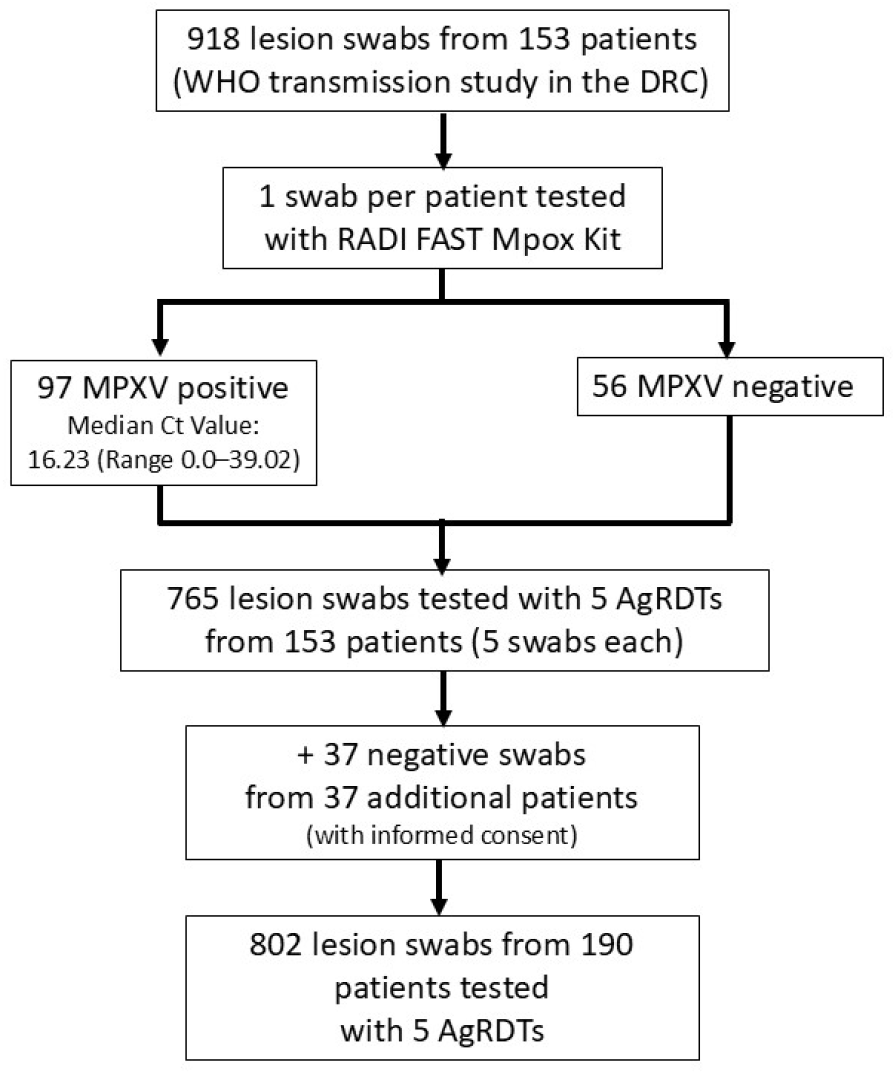
Diagnostic testing of mpox lesion swabs with antigen-based rapid diagnostic tests, Kinshasa, the Democratic Republic of the Congo.

The demographic and clinical characteristics of the participants are shown in Table 1.

**Table 1.** Age and sex characteristics of MPXV positive and negative patients from DRC included in the evaluation.

| Characteristics | MPXV Positive, n/N (%) | MPXV Negative n/N (%) | Overall n/N (%) |
| --- | --- | --- | --- |
| **Age group** |  |  |  |
| **0-14 years old** | 14/37 (37.8) | 23/37 (62.2) | 37/37 (100) |
| **15-59 years old** | 83/152 (54.6) | 69/152 (45.4) | 152/152 (100) |
| **60+ years old** | 0/1 (0.0) | 1/1 (100) | 1/1 (100) |
| **Sex** |  |  |  |
| **Female** | 37/75 (49.3) | 38/75 (50.7) | 75/75 (100) |
| **Male** | 60/115 (52.2) | 55/115 (47.8) | 115/115 (100) |
| **M/F ratio** | - | - | 115/190 (60.5%) |

Diagnostic performance against the reference test showed considerable variability among the five AgRDTs. Among the assays, Monkeypox Virus Ag Test Kit from Guandong Wesail Biotech demonstrated the best overall balance, with a sensitivity of 77.3% (95% CI: 68.0-84.5) and a specificity of 93.5% (95% CI: 86.6-97.0). Monkeypox Antigen Test Cassette from Hangzhou Testsea Biotechnology also performed relatively well, achieving a sensitivity of 72.2% (95% CI: 62.5-80.1) and a specificity of 93.5% (95% CI: 86.6-97.0). Monkeypox Virus Antigen Rapid test from Beijing Hotgen Biotech and Monkeypox Virus Antigen Rapid Test Kit from Contipharma showed moderate sensitivity (59.8% [95% CI 49.8-69.0] and 50.5% [95% CI 40.7-60.3], respectively) despite maintaining high specificity (>95%). NG-Test Monkeypox virus from NG Biotech performed poorest, with a sensitivity of 39.2% (95%CI: 30.1-49.1), although its specificity remained high (96.8% [95%CI: 90.9-98.9], Figure 2).

**Figure 2:**
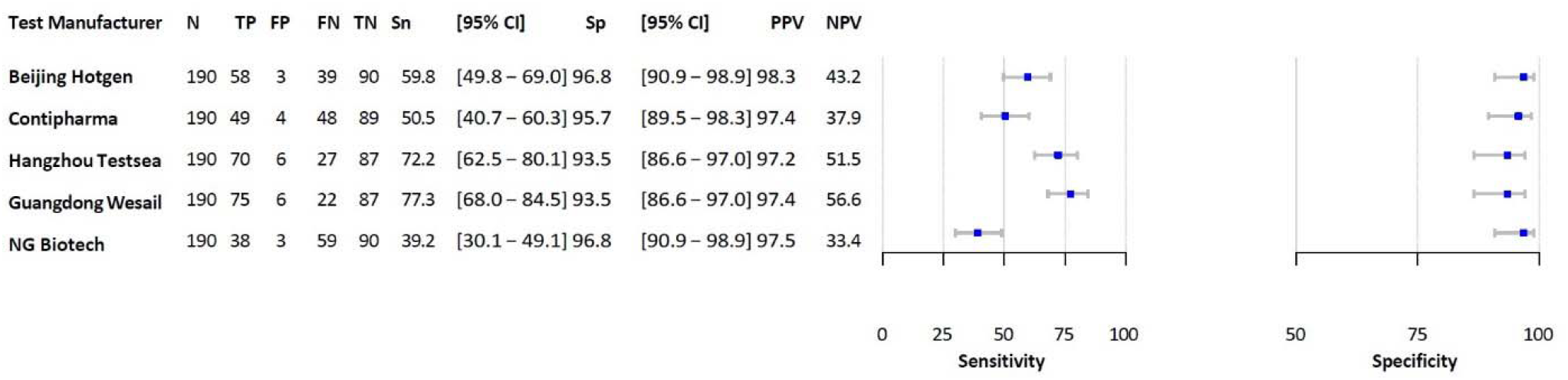
Performance characteristics of five AgRDTs (index tests) compared with PCR testing (reference test). TP: true positive; FP: false positive; FN: False negative; TN: true negative; Sn: sensitivity; Sp: specificity; PPV: positive4 predictive value; NPV: negative predictive value

#### Secondary analyses

A comparison of the sensitivity for clade Ia and clade Ib (Figure 3) highlights variations in the diagnostic sensitivity of the five tests. For clade Ia (N=70), Monkeypox Virus Ag Test Kit from Guandong Wesail Biotech showed the highest sensitivity at 70.0% (95% CI: 58.5–79.5), followed by Monkeypox Antigen Test Cassette from Hangzhou Testsea Biotechnology at 64.3% (52.6–74.5), Monkeypox Virus Antigen Rapid test from Beijing Hotgen Biotech at 54.3% (42.7–65.4), Monkeypox Virus Antigen Rapid Test Kit from Contipharma at 45.7% (34.6–57.3), and NG-Test Monkeypox virus from NG Biotech at 37.1% (26.8– 48.9). In contrast, clade Ib sensitivity (N=27) was markedly improved for most tests, although sample size was much smaller. Monkeypox Virus Ag Test Kit from Guandong Wesail Biotech again performed best with a sensitivity of 96.3% (81.7–99.3), closely followed by Monkeypox Antigen Test Cassette from Hangzhou Testsea Biotechnology at 92.6% (76.6–97.9). Monkeypox Virus Antigen Rapid test from Beijing Hotgen Biotech and Monkeypox Virus Antigen Rapid Test Kit from Contipharma achieved moderate sensitivities of 74.1% (95% CI 55.3–86.8) and 63.0% (95% CI 44.2–78.5), respectively, while NG-Test Monkeypox virus from NG Biotech remained the least sensitive at 44.4% (95% CI 27.6–62.7). Overall, both datasets consistently identify Guangdong Wesail Biotech and Hangzhou Testsea Biotechnology as the most reliable test manufacturers, while NG Biotech assay shows persistently lower sensitivity, indicating substantial variability in test performance under different conditions or populations.

**Figure 3:**
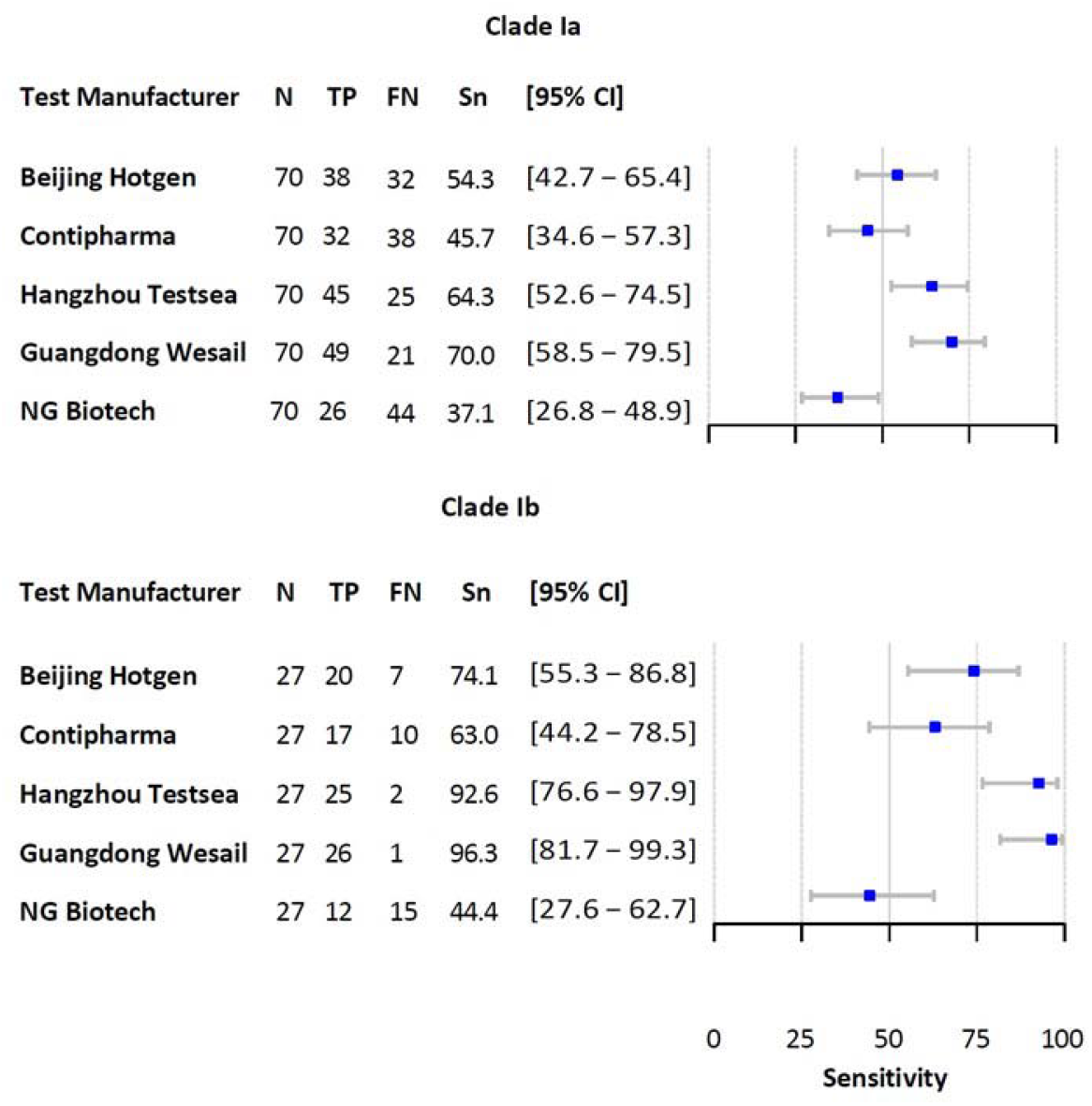
Performance characteristics of five AgRDTs (index tests) compared with PCR testing (reference test) by MPXV clade. TP: true positive; FN: False negative; Sn: sensitivity.

When comparing sensitivity by Ct values, the tests demonstrated higher sensitivity at lower Ct values and a general decline as Ct increased, indicating a strong dependence on viral load. Three test manufacturers (Beijing Hotgen Biotech, Hangzhou Testsea Biotechnology, and Guangdong Wesail Biotech) maintained the expected average sensitivity up to the third Ct quartile, whereas Contipharma and NG Biotech showed a more pronounced decline from earlier quartiles. In the highest Ct quartile (>24.38), sensitivity dropped sharply for all tests, with point estimates generally below 30%, indicating limited ability to detect low-viral-load infections (Figure 4).

**Figure 4:**
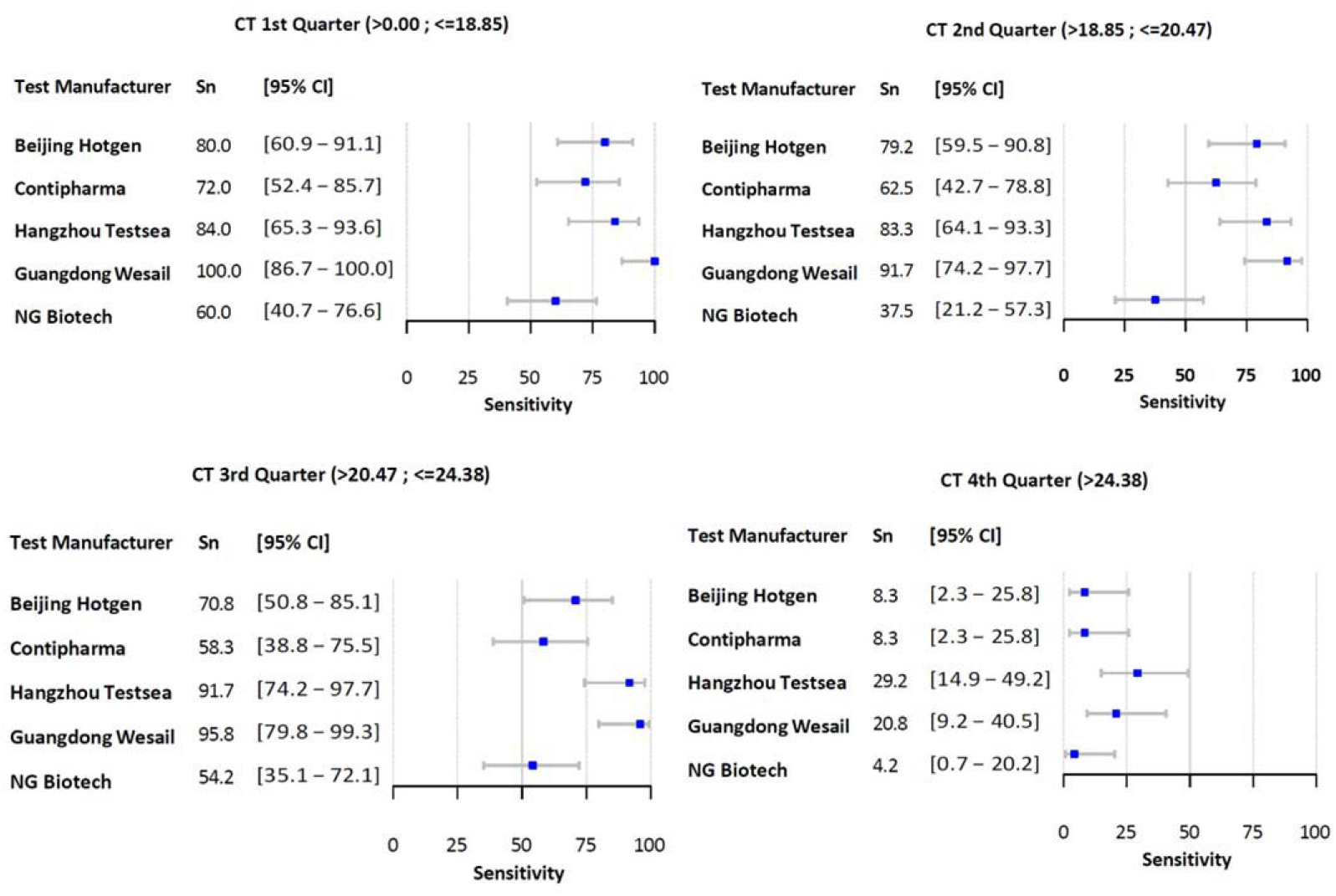
Performance characteristics of five AgRDTs (index tests) compared with PCR testing (reference test) by Ct value quarter. Sn: sensitivity.

The sensitivity of all five diagnostic tests varied notably between children (≤15 years) and adults (>15 years, Supplementary Figure 1). Across all tests, sensitivities are consistently higher in adults compared to children. The test from Guangdong Wesail Biotech shows the highest performance in both groups, with 56.2% (95% CI 33.2–76.9) in children and 81.5% (95% CI 71.7–88.4) in adults, followed by the one from Hangzhou Testsea Biotechnology at 50.0% (95% CI 28.0–72.0) in children and 76.5% (95% CI 66.2– 84.4) in adults. Those from Beijing Hotgen Biotech and Contipharma show moderate sensitivities, with both improving substantially in adults. The test from NG Biotech demonstrates the lowest sensitivity overall, with minimal improvement from 37.5% (95% CI 18.5–61.4) in children to 39.5% (95% CI 29.6– 50.4) in adults.

### Analytical evaluation

The analytical testing with culture virus was conducted to orient on sensitivity for Clade Ib using the same five Ag-RDTs that were used with patient samples. None of the AgRDTs detected viral dilutions with a Ct > 30, equivalent to concentrations lower than 10,000 DNA copies per ml or 1,000 DNA copies applied per test (Table S1). Infectious viral particles were present in dilutions with a limit of detection (LOD) down to 10^3^ copies/ml, indicating that AgRDTs would miss a proportion of samples with infectious titres. There were no significant differences in test sensitivity between the five Ag-RDTs; however, the most sensitive test (Guangdong Wesail Biotech RDT, LOD: 3.55×10^5^ DNA copies/mL) detected one log more DNA copies than the least sensitive test (Beijing Hotgen Biotech RDT, LOD: 3.81×10^6^ DNA copies/mL).

## Discussion

This evaluation of five mpox AgRDTs provides insight into their analytical and clinical performance in settings where MPXV clade I is endemic and access to molecular diagnostics is limited. While none of the tests fully met WHO TPP criteria, two assays—Hangzhou Testsea Biotechnology and Guangdong Wesail Biotech—met the ≥70% sensitivity and approached specificity (93.5%) benchmarks set for the study following the first evaluations of AgRDTs, which had resulted in disappointing findings, ^23^ and their sensitivity increased markedly for MPXV clade Ib samples (93–96%).

High specificity was observed for three assays (Beijing Hotgen Biotech, Contipharma, and NG Biotech), all meeting or exceeding the ≥95% threshold. In hyperendemic settings without routine PCR access, high specificity is critical to limit false positives and unnecessary isolation or inappropriate treatment. However, the resulting low negative predictive value indicates that negative AgRDT results cannot reliably rule out mpox, confirming that these assays cannot replace molecular diagnostics to confirm a negative result.

One assay demonstrated the lowest analytical limit of detection and the highest clinical sensitivity, suggesting internal consistency between analytical and clinical performance. Despite their limitations, some AgRDTs may still have a role as screening tools in settings where access to PCR testing is unavailable. In remote or resource-limited environments, these assays, which do not require the cold chain, could be used for preliminary case identification or triage, rather than as definitive diagnostic tools. Their deployment should be informed by local epidemiology, with circulating variants ideally characterized through periodic sentinel PCR testing or sequencing where feasible, to support more targeted and appropriate use. WHO is currently working to update recommendations on mpox testing, which could include potential use of AgRDTs.

Differences in sensitivity compared with previous evaluations of the NG-Test Monkeypox virus^18^ may reflect variation in sampling procedures, circulating clades, or other confounders. In this study, most samples were MPXV clade Ia, with substantially lower sensitivity than for clade Ib.

No defining amino acid differences were identified in the A29 protein between clades Ia and Ib. However, minor sequence variations can alter epitope conformation and antibody binding, particularly on exposed surface antigens, potentially explaining clade-specific performance differences and underscoring the importance of antigen selection in assay design.^24^

Reduced sensitivity at high Ct values was expected, reflecting low viral loads. Given the complexity of MPXV antigens, AgRDTs based on monoclonal antibodies are inherently less sensitive than molecular assays and require sufficient viral material for detection.

Age-related differences in sensitivity, also reported by Bakamutumaho et al., ^18^ may reflect procedural rather than biological factors. In paediatric patients, discomfort or limited cooperation may result in less vigorous swabbing, thereby reducing diagnostic sensitivity. Guangdong Wesail and Hangzhou Testsea showed the most consistent performance across age groups.

## Limitations

This retrospective study was based on archived dry lesion swabs collected over the preceding six months, which were stored at −80 °C shortly after collection and remained frozen until analysis; point-of-care testing was not conducted at the time of specimen collection. The restriction of sampling to Kinshasa Province and the predominance of MPXV clade I may limit the generalizability of the findings, particularly to rural or epidemiologically distinct settings. In addition, analytical sensitivity assessments were performed using clade Ib virus stocks only, although previous studies suggest minimal clade-dependent differences in analytical performance.^25^

Use of different reference standards for analytical and clinical evaluations, and limited number of replicates for the analytical sensitivity work introduce additional uncertainty. The modest sample size resulted in wide confidence intervals, particularly for subgroup analyses. Few high-Ct samples were included, and analytical data indicate that such specimens are unlikely to yield positive results with the AgRDTs assessed in this study.

To comply with manufacturers’ instructions and avoid rehydrating dry swabs with VTM – which was shown to interfere with AgRDTs ^16^-or other external liquid, an approach using multiple lesion samples from the same individual was adopted; however, this introduces a potential source of bias, as testing different samples with different AgRDTs is not equivalent to testing aliquots of a single, homogenized sample, and inherent intra-individual and sampling variability may affect assay performance independently of test characteristics.

## Conclusion

Overall, these findings clarify that mpox AgRDTs should be viewed as context-dependent public health tools rather than substitutes for molecular diagnostics. In hyperendemic, resource-limited settings, selected higher-performing assays can support earlier case identification, triage, and outbreak signal detection, thereby enabling faster isolation and response while conserving PCR capacity for confirmatory testing and surveillance. Critically, this evaluation highlights that antigen selection and clade-specific performance must inform procurement, policy, and future test development, reinforcing the need for adaptive diagnostics aligned with local epidemiology rather than universal deployment. Further studies are needed to better understand the performance of these tests and better define use cases. A multi country field-based validation of most promising AgRDTs is ongoing and WHO is updating recommendations on the possible use of AgRDTs in mpox surveillance and outbreak response.

## Data Availability

All data produced in the present study are available upon reasonable request to the authors

## Author contributions

The study was conceived and designed by LS, DME, EA, HFC, and IE. Laboratory work was conducted by EIN, RL, MMuk, MMul, CE, KA with supervision by HKM, IE and PMK. Data collections were conducted by EIN, RL, CE, MMuk, MMul, KA. Data analysis and interpretation were conducted by LS, HFC, CE, IE, HK, EA, DMB, JCMC, EKL, ON, RL, NG, RS, RFL, DMB, RF, TWB and PMK. The initial manuscript was prepared by LS. All authors edited and approved the final manuscript.

## Declaration of Competing Interest

The authors declare that they have no known competing financial interests or personal relationships that could have appeared to influence the work reported in this paper.

## Funding

WHO

## Disclaimer

The views expressed here are solely those of the authors and do not necessarily reflect the views of their respective employers.

## Acknowledgements

We acknowledge the participants for volunteering for this study. We thank all the research nurses and epidemiologists of the transmission study for supporting with the sample collection and recruitment.

---

**Supplementary Table 1:** List of manufacturers and respective index tests shortlisted for the evaluation.

| Test name | Test manufacturer (Country) | Claimed limit of detection | Regulatory approval | Claimed clinical sensitivity | Claimed clinical specificity |
| --- | --- | --- | --- | --- | --- |
| Monkeypox Virus Antigen Rapid test | Beijing Hotgen Biotech (China) | 100 PFU/mL | CE-IVDD, UK MHRA | 99.6% | 99.2% |
| Monkeypox Virus Antigen Rapid Test Kit | Contipharma (Belgium) | 250 PFU/mL | CE-IVDD | 95.5% | 99.5% |
| TestSea Monkeypox Antigen Test Cassette | Hangzhou Testsea Biotechnology (China) | 1000 TCID<sub>50</sub>/mL | CE-IVDD | 98.0% | 99.6% |
| Monkeypox Virus Ag Test Kit | Guangdong Wesail Biotech (China) | 6.25 PFU/mL | CE-IVDD, UK MHRA | 93.0% | 99.0% |
| NG-Test® Monkeypox virus | NG Biotech (France) | 32,000 TCID<sub>50</sub>/mL | RUO | - | - |

**Supplementary Table 2.** Analytical testing results with clade Ib viral dilutions for the five MPXV antigen rapid tests.

| Dilution | Ct (ref PCR) | Concentration (DNAC/mL) | NG Biotech |  |  | Beijing Hotgen |  |  | Contipharma |  |  | Hangzhou Testsea |  |  | Guangdong Wesail |  |  |
| --- | --- | --- | --- | --- | --- | --- | --- | --- | --- | --- | --- | --- | --- | --- | --- | --- | --- |
|  |  |  | Rep 1 | Rep 2 | Rep 3 | Rep 1 | Rep 2 | Rep 3 | Rep 1 | Rep 2 | Rep 3 | Rep 1 | Rep 2 | Rep 3 | Rep 1 | Rep 2 | Rep 3 |
| 1 | 23.09 | 1.21E+07 | + | + | + | + | + | + | + | + | + | + | + | + | + | + | + |
| 2 | 24.78 | 3.81E+06 | + | + | + | + | + | + | + | + | + | + | + | + | + | + | + |
| 3 | 26.37 | 1.31E+06 | + | + | + | - | - | - | + | + | + | + | + | + | + | + | + |
| 4 | 28.30 | 3.55E+05 | - | - | - | - | - | - | - | + | - | - | - | - | + | + | + |
| 5 | 30.31 | 1.07E+05 | - | - | - | - | - | - | - | - | - | - | - | - | - | - | - |
| 6 | 32.51 | 2.19E+04 | - | - | - | - | - | - | - | - | - | - | - | - | - | - | - |
| 7 | 33.77 | 9.45E+03 | - | - | - | - | - | - | - | - | - | - | - | - | - | - | - |
| Neg | N.A. | N.A. | - | - | - | - | - | - | - | - | - | - | - | - | - | - | - |

**Supplementary Figure 1:**
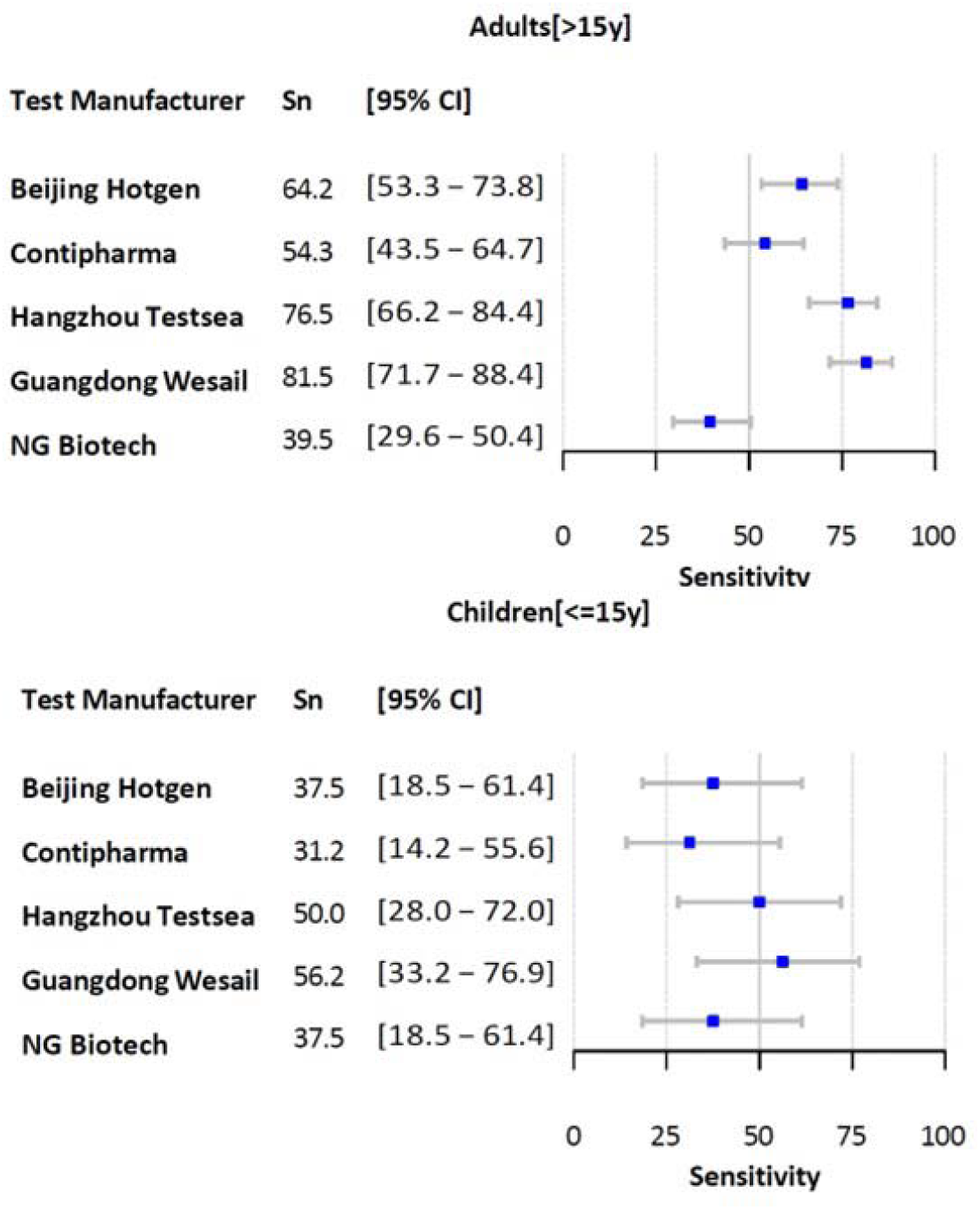
Performance characteristics of five AgRDTs (index tests) compared with PCR testing (reference test). Sn: sensitivity.

## Notes

### Competing Interest Statement

The authors have declared no competing interest.

