## Supplementary figures and images for "Diagnostic accuracy of five mpox lateral flow assays for antigen detection, the Democratic Republic of the Congo and Switzerland"

### Supplementary Figure 1.jpg

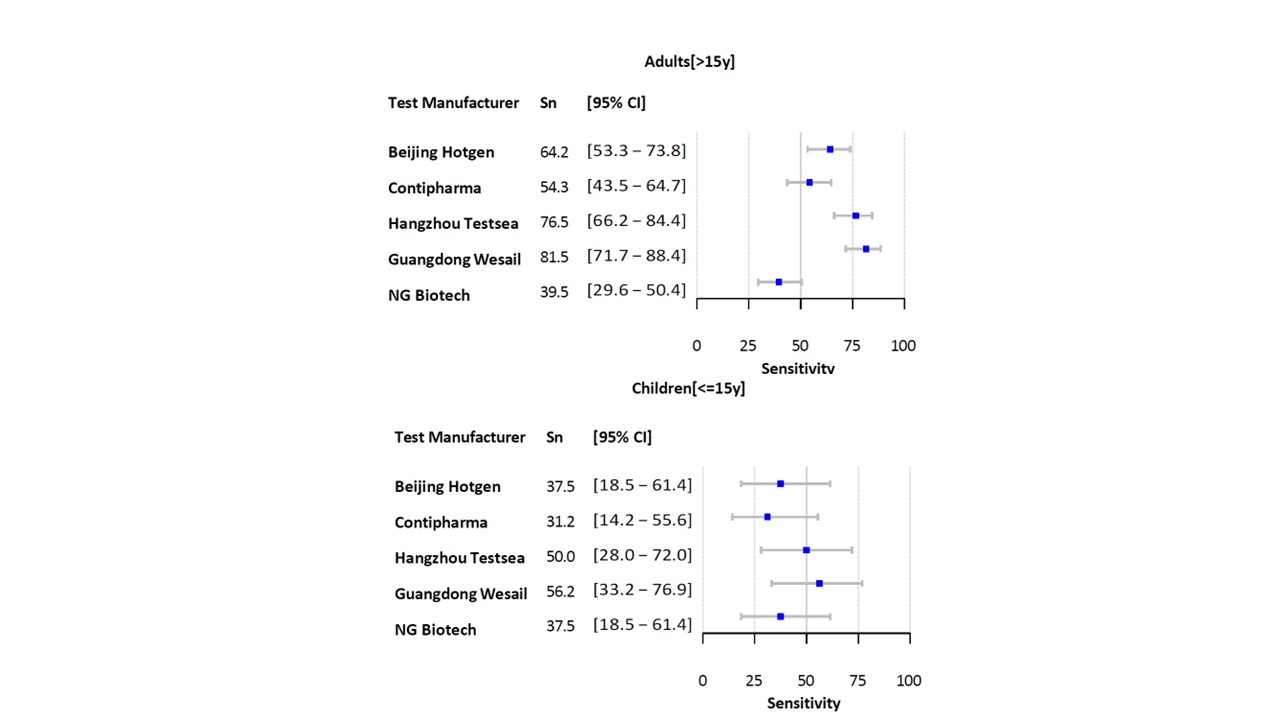
